# Visual Display Interface (VDI): A MATLAB Software Library For Simulating and Processing In-Vivo Magnetic Resonance Spectroscopy and Spectroscopic Imaging Data

**DOI:** 10.1101/2023.08.31.23294888

**Authors:** Yiling Liu, Georg Oeltzschner, Itamar Ronen, Rita Schmidt, Amir Seginer, Ivan Kirov, Dan Wu, Zhiyong Zhang, Assaf Tal

**Author notes:** **Corresponding Author:** Assaf Tal, PhD, Department of Chemical and Biological Physics, Weizmann Institute of Science, 234 Herzel St., Rehovot 7610001, Israel.

## Abstract

We describe a new comprehensive and open-source MATLAB library, called the Visual Display Interface (VDI), designed to process, model, quantify and visualize magnetic resonance spectroscopy (MRS) and spectroscopic imaging (MRSI) data. The library focuses on three major strengths: promoting reproducible research by creating comprehensive data logging and reporting and identifying outlier datasets; seamlessly combining spectral and spatial processing of both single and multivoxel data; and offering a modern, object-oriented design. VDI handles a wide range of common tasks, including spectral and spatial transforms, built-in spectral fitting, absolute quantification, and running density-matrix simulations for coupled and uncoupled spin systems to generate appropriate basis sets and aid in sequence design. VDI interfaces with the Statistical Parameteric Mapping (SPM) toolbox to carry out tissue segmentation, calculate tissue fractions within voxels, derive mean metabolite values from regions of interest defined by anatomical or functional atlases, and perform linear regression for global white matter and gray matter metabolite concentrations. The library’s workings are demonstrated for two tasks: (1) Pre-processing, fitting and analysis of single-voxel proton MRS data from healthy volunteers; and (2) Extracting region-specific metabolite concentrations from spectroscopic imaging data based on an existing cortical atlas in MNI space, and calculating average gray and white matter global concentrations using linear regression.

## Introduction

The analysis of Magnetic Resonance Spectroscopy (MRS) and spectroscopic imaging (MRSI) data involves the manipulation of spatial, temporal and spectral degrees of freedom. Such analyses might include registering averages, carrying out phase corrections, fitting spectra, quantifying metabolite concentrations, calculating the mean value of a metabolite inside an anatomical region of interest, calculating fractions of gray matter and white matter tissue within a spectroscopic voxel, producing metabolic maps that are properly aligned with high resolution structural images. These require unique and highly-specialized computational tools. Over the past decades, several packages have been proposed for handling some of these aspects, including FID-A^1^, FiTAID^2^, FSL-MRS^3^, Gannet^4^, INSPECTOR^5^, jMRUI^6^, LCModel^7,8^, MRSCloud^9^, Osprey^10^, SPANT^11^, TARQUIN^12^ and VESPA^13^. Some of these freely available packages can handle spectral fitting and quantification, others preprocessing. A few are equally geared towards single and multivoxel data. Some, but not all, provide spin simulation capabilities.

Here, we describe a new set of libraries, written in MATLAB (The Mathworks, Natick, MA, USA) and compatible with releases 2018b onward, called the Visual Display Interface (VDI). VDI was specifically written to enhance the reproducibility of MRS research by focusing on three key quality-of-life areas: Provide users a modern, object-oriented interface for handling MRS data which promotes writing self-documenting code; Seamlessly combine the handling of multiresolution MRS and MRSI data, by providing a single coherent framework to oversee all spectral and spatial data processing; And provide in-depth HTML reporting and analysis report-generating capabilities for detecting outliers and summarizing data statistics which is specifically suited for large datasets with multiple subjects. More specifically, VDI is:

1. Freely and widely accessible: VDI is open-source and freely available for download (via www.vdisoftware.net), and is implemented in MATLAB, one of the most widely-used scientific computing packages in academia.
2. Extensively documented: The VDI libraries are extensively documented. They were written to anticipate a wide range of user inputs. An extensive set of examples is provided in a separate subdirectory titled “examples”.
3. Report-Generating: Preprocessing, fitting, an simulations all produce HTML reports which provide numerical and graphical feedback, as well as try to identify corrupted or outlier data. This is done both at the single-subject and group levels. All figures which appear herein were pasted directly from the generated reports, with no or minimal modifications.
4. Accessible to beginners: By adopting an object-oriented architecture, VDI provides users with clear, robust and accessible command-line tools that produce self-documenting and succinct code. Most parameters are automatically determined by the routines without the need for any user intervention.
5. Extendible by experts: The object-oriented structure of VDI provides a rich and elegant framework for producing highly complex scripts capable of handling diverse datasets.
6. Integrated with SPM: The Statistical Parametric Mapping package (https://www.fil.ion.ucl.ac.uk/spm/) offers powerful MATLAB-based image segmentation and processing capabilities. VDI can call several SPM routines directly to handle a variety of tasks, from segmenting T_1_-weighted images into gray matter, white matter and cerebrospinal fluid maps, to the calculation of metabolite statistics within atlas-based predefined regions of interest.
7. Capable of handling multimodal, multiresolution data: VDI has been written to handle multidimensional (2D and 3D) spectroscopic imaging data, and can operate efficiently on such data. Some of its capabilities include masking; calculating tissue fractions within voxels; tissue-type regression; and calculation of descriptive statistics within regions of interest. Users can provide their own masks in DICOM of NifTI format, or employ SPM to generate such masks.
8. Interactive and visual: The library offers interactive tools for displaying and interacting with both spectral and spatial data dimensions in real-time. Furthermore, the library produces and summarizes several of its data structures in table format, and offers powerful tools for plotting and exploring such tabular data using a variety of plot types.

VDI is already in use by several groups and underlies the processing of data in multiple publications over the past several years^14–20^. To facilitate its widespread dissemination, the current manuscript describes the general philosophy behind the framework, and its main classes and the way they interact among themselves. Two examples of usage of the library are presented in the Methods section, and highlight its versatility: In the first, SemiLASER single-voxel MRS data are loaded, preprocessed, fitted, quantified (in terms of absolute concentrations) and visualized. In the second, global linear regression is carried out on a 32×32 2D MRSI supra-ventricular slice from a healthy volunteer to obtain average white matter and gray matter tissue metabolite concentrations^21–23^.

## Theory

### Data Structures for Single and Multivoxel Data

VDI employs an object oriented approach to handling spatial data (Fig. 1), where data and methods are grouped into classes. Inheritance, in the object-oriented sense, is implemented wherever possible – that is, some classes retain and extend the capabilities of others – and indicated by black arrows. At its root is the class VDIVolume, which describes a general 3D oriented volume in space, with position, size and orientation metadata. These properties are given relative to some common reference frame, which for most applications coincides with that of the magnet’s frame of reference, with its center at the center of the gradient system. `VDIVolumeMatrix` inherits from `VDIVolume` and enriches it by adding the ability to segment `VDIVolume` into voxels. However, `VDIVolumeMatrix` is rarely used itself. Rather, its children, which can also associate data with the voxels, are much more useful: `VDIImage`, which is used to store 3D data, such as structural MRI images or metabolite maps; and `VDIImageND`, which is used to store n-dimensional data (in the `data` property), such as spectroscopic imaging or BOLD-fMRI data. Thus, `VDIImageND` is the most general class for storing and manipulating spectral-spatial-temporal data, such as functional MRS^16,17,24–26^, hyperpolarized ^13^C-MRS^27,28^, deuterium metabolic imaging data^29–31^, spectroscopic fingerprinting^18,32,33^. However, `VDIImage` offers greater flexibility, a smaller footprint and specialized methods specific to 3D imaging data. A third important class that derives from `VDIVolume` is `VDIShape`, which is used to describe three-dimensional boxes and ellipsoids, which can be used, e.g., to define regions of interest for processing, or the position of a voxel.

**Figure 1.**
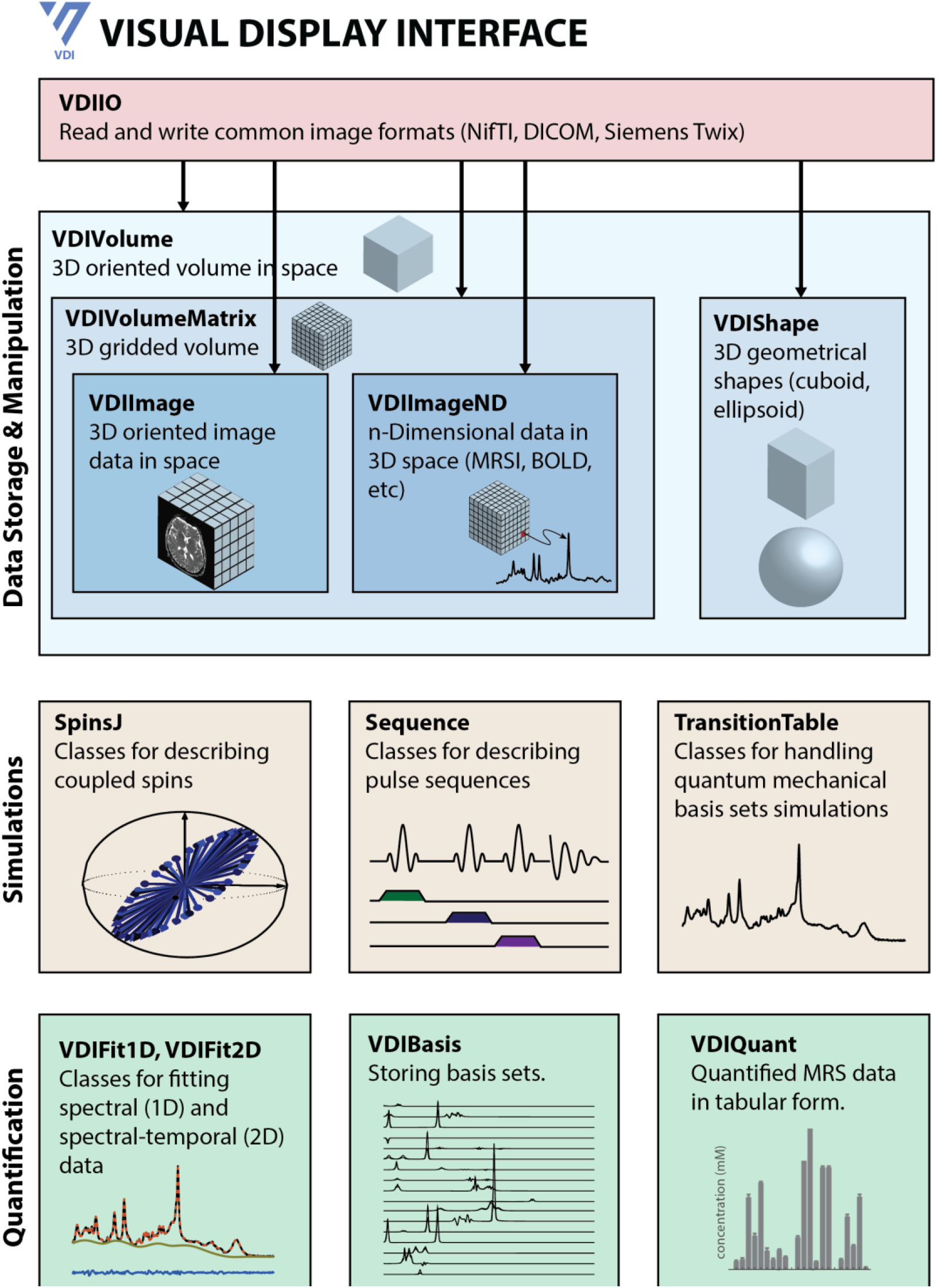
The major classes in the VDI framework, and their various relationships. Arrows indicate class inheritance, in the object-oriented sense. The main classes for storing and manipulating MRS and MRSI data are shown in the top (blue) row. Classes for carrying out quantum mechanical spin simulations are shown in the second row (yellow). The final row details major classes involved in fitting and modeling spectral data.

The use of object oriented design and operator overloading (namely, the redefinition of default operators such as addition or element-wise multiplication) allows for the seamless manipulation of spatial-spectral datasets, even with different orientations, positions and spatial resolutions. Consider, for example, the following code, which zeros out voxels in a low-resolution `VDIImage` metabolic map of n-acetyl-aspartate (NAA) outside the posterior cingulate gyrus (PCG), using a high resolution binary mask:

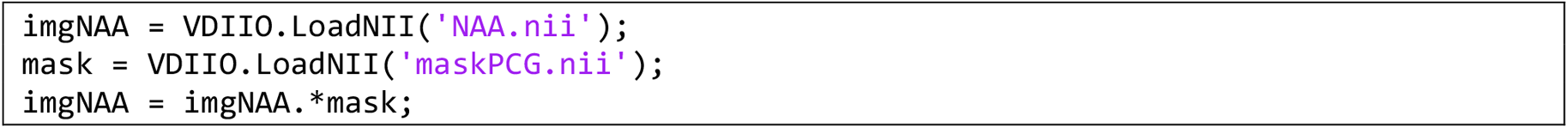

This code uses VDIIO to load the NAA metabolic map and PCG mask – both of which are `VDIImage` objects – and uses the fact that the .* operator is overloaded for the `VDIImage` class. It interpolates the high resolution mask of the PCG onto the spatial grid of the NAA image before carrying out the multiplication. All interpolations within VDI are nearest-neighbors: the value of each voxel in the interpolated image equals that of the voxel in the high-resolution image closest to it (center-to-center). VDI classes have overloaded several common MATLAB operators, including element-wise multiplication, addition, subtraction and element-wise division.

Spectroscopic data, whether single or multivoxel, are stored as `VDIImageND` objects. These objects offer extensive capabilities for preprocessing, visualizing and manipulating such data (as shown in the Methods section below). `VDIImageND` objects are initialized with an n-dimensional array of data and appropriate labeling of each dimension type, space, unit and axis. For example, 6-dimensional spatial-spectral array `data` from multiple coils with dimensions (x, y, z, time, averages, coil) can be initialized as follows:

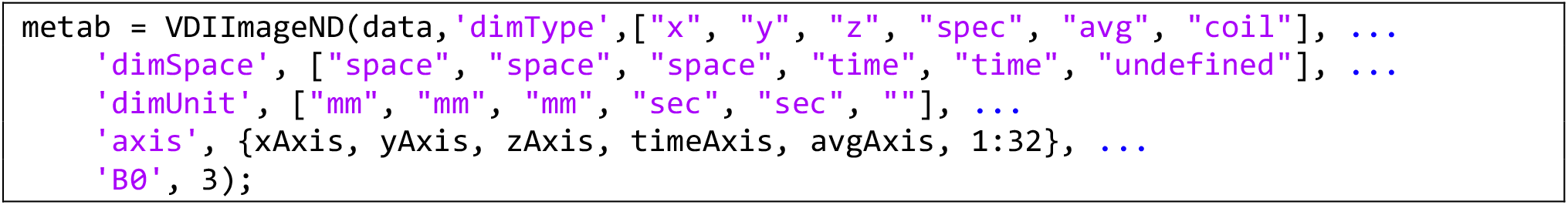

`xAxis`, `yAxis`, `zAxis` and `timeAxis` are all vectors (defined elsewhere), indicating the relevant axes for the data along each of its dimensions (e.g., `yAxis` is in millimeters, and has the same number of elements as data along its second dimension).

### Preprocessing of Data

Preprocessing refers to several standard steps in which spectral and spatial data is modified prior to spectral fitting. Methods for carrying out common preprocessing steps are implemented for `VDIImageND` objects. For example, coil addition, spectral registration, and automated phasing can all be carried out with a few commands:

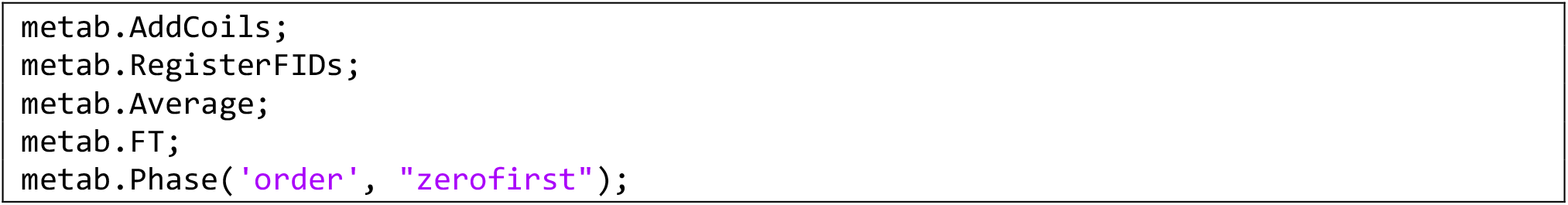

These methods obtain most required metadata from the `VDIImageND` object itself and therefore require little additional input, but can also be tailored with additional pairs of variable names and values. For example, phasing only corrects zero order distortions. However, in the above example, both zero and first order phasing are requested by setting the input parameter `order` in the `Phase` method to `zerofirst`. Some of the more commonly used preprocessing methods implemented for `VDIImageND` are described in Table 1, alongside the algorithms they implement.

**Table 1.** Several common VDIImageND methods used to preprocess data.

| Action | Method | Description |
| --- | --- | --- |
| Coil Combination | AddCoils | Singular-Value decomposition coil addition<sup>34,35</sup>, modeling the data as a linear combination of signals $s_j$ which differ up to a complex constant $a_j$: $s(t) = \sum_j s_j a_j$. By default, addition coefficients are proportional to the signal intensity. If a reference noise dataset is provided, coefficients become proportional to the SNR. |
| Spectral Registration | RegisterFIDs | An spectral-alignment and zero-order phasing algorithm following Mikkelsen et. al.<sup>36</sup>, in which FIDs are sorted according to similarity and then iteratively weighted according to similarity, phased and aligned. |
| Averaging | Average | Allows for simple or complex averaging patterns. For example: Average('dim', "set", 'pattern', [1 2], 'coeffs', [1 -1]) subtracts consecutive averages, as done in MEGAPRESS. More complex phase cycling reconstruction schemes can easily be implemented this way, by setting coeffs accordingly. |
| Apodization | Apodize | Gaussian (default) or Lorentzian apodization. Can be equally applied along spectral and spatial dimensions. |
| Zero-Filling | ZF | Zero-filling, either one-sided (default) or symmetrical. |
| Automatic Phasing | Phase | Minimum-entropy based automatic phasing for either zero and/or first order phase distortions<sup>37</sup>. |
| Manual Phasing | N/A | Manual zero-order phasing can be applied by using the overloaded .* operator. For example, typing: myData.*exp(1i*pi) |
| Fourier Transform | FT | Fourier Transforms are handled automatically based on the dimension type (spectroscopic, spatial), and space (freq, time for the spectral dimension; space or kspace for spatial dimensions). |

**Table 2.** VDIImageND methods for quality assurance of MRS and MRSI data.

| Method | Description |
| --- | --- |
| EstimateFWHM | Estimate water full-width at half maximum (can also be used for any single, well-resolved peak). |
| EstimateLipidContamination | Estimate spectral lipid contamination, defined as the ratio of the maximal intensity of the signal in the [0.3, 1.7] ppm range to the maximum of the NAA signal (in the [1.85, 2.15] ppm range). |
| EstimateSNR | Estimate the signal-to-noise ratio of the NAA peak to the real part of the noise. The noise is either estimated from a reference noise dataset, or the [7, 9] ppm range in the spectral dataset. Noise is linearly detrended by default. |
| DetectOutlierSpectra | Detects outlier spectra *across subjects*, defined via their $L_2$ distance from the median spectrum. Only works on the arrays of VDIImageND objects. By default, this method operates voxel-by-voxel; normalizes all spectra by their individual $L_2$-norms prior to commencing outlier detection; takes the absolute value of the spectrum; operates on the [0.2,4.2] ppm range. Outliers are defined as spectra with $L_2$-norms more than three scaled absolute median deviations from the median. |

A second powerful set of methods allows the user to extract quality assurance metrics for their spectroscopic data, such as linewidth, SNR, and FWHM. Default behaviors, which can be modified via optional parameter inputs.

Almost all `VDIImageND` methods offer the ability to produce detailed HTML reports of their actions, by accepting a `VDILog` object, as demonstrated by the following snippet:

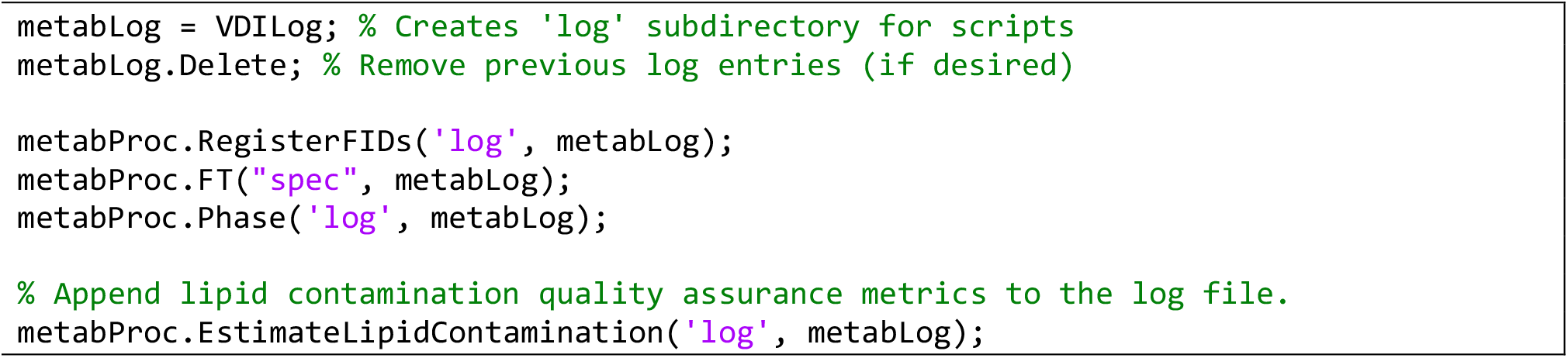

### Spin Simulations

VDI is provided with an independent sub-library called SpinTool, hosted on a separate path, which contains code for simulating the behavior of both uncoupled and coupled spin systems (`SpinsJ` class) in response to pulse sequences (`Sequence` class). It does so by solving the full quantum-mechanical Liouville equations of motion, without relaxation. Spins are initialized in a straightforward manner. For example, creating a spin ensemble with NAA, creatine (Cr) and phosphocreatine (PCr) can be done with two lines:

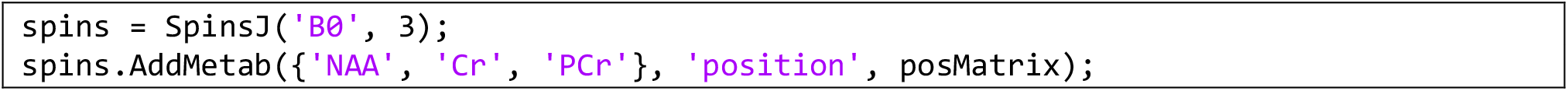

`posMatrix` is a N×3 matrix, with each row corresponding to an [x,y,z] position at which to place a metabolite instance, allowing for arbitrary 3D distributions (e.g., `posMatrix=[0,0,0]` will place a single copy of each metabolite at the center of the magnet system).

Pulse sequences can be defined easily using a user-friendly syntax. For example, a spin-echo sequence using idealized hard pulses can be coded within a single line, and applied to the previously-defined spin structure with an additional line of code. A matrix containing the time-domain simulated basis function for each metabolite is created in a third line:

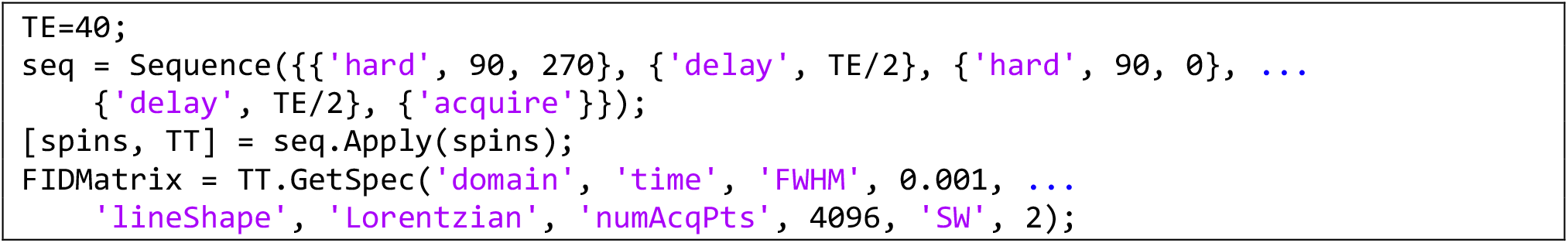

More complex sequences, containing arbitrary pulse waveforms and phase cycling can be created just as easily. This sequence can be easily visualized using the `Plot` method in the `Sequence` class (i.e. by typing `seq.Plot`). The `Apply` method returns the spins following the sequence. Acquired spectral data `TT` is stored in the `TransitionTable` class; each `TransitionTable` object described the spectral pattern associated with a particular metabolite, in terms of the frequencies, amplitudes and phases associated with each resonance. These can be convolved with appropriate lineshape functions – e.g. Lorentzians or Gaussians – to generate either free induction decays in the time domain, or spectra in the frequency domain. This is done using the `GetSpec` method, which accepts as inputs multiple parameters, such as the full-width-at-half-maximum (FWHM) and spectral width (SW) for shaping the spectral acquisition parameters.

### Fitting, Spectral Modeling and Absolute Quantification

The `Sequence` and `SpinsJ` classes offer the necessary machinery for generating basis sets, which are essential for spectral fitting. Basis sets are handled via the `VDIBasis` class. Once a TransitionTable object has been obtained via simulations, it can be converted into a basis set, and exported to a format consistent with the popular LCModel fitting package^7,8^, within a few lines of code:

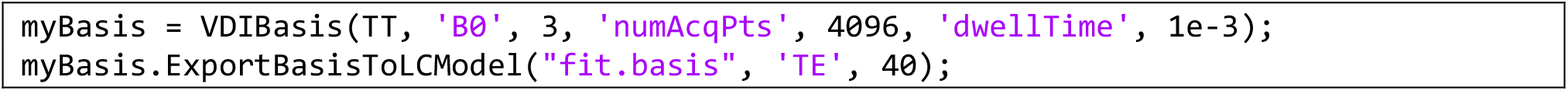

LCModel has recently been released to the public domain (https://github.com/schorschinho/lcmodel) and offers a free, robust, widely validated fitting package; its precompiled binaries need only be added to the MATLAB path. Spectroscopic data in `VDIImageND` objects – whether single or multivoxel – can then be directly fit using the `FitLCModel` method.

VDI contains two additional classes, distinct from LCModel, which implement least squares fitting of spectroscopic data, appropriately named `VDIFit1D` and `VDIFit2D`. The first can be directly used as a replacement for LCModel for fitting one-dimensional spectroscopic data. `VDIFit2D` has been specifically created to fit spectral-temporal two-dimensional datasets, where temporal correlations exist between successive transients. These approaches have been suggested (and subsequently also implemented in FSL-MRS^38^) to substantially improve fitting precision^39^ by providing additional temporal models (i.e. constraints) for the time evolution of metabolite amplitudes, frequencies, phases or lineshape parameters^2,3^.

All spectral fitting routines generate tabular data which is stored in objects belonging to the `VDIQuant` class. The `metabTable` property of a `VDIQuant` object contains a table in which each metabolite from each region and volunteer occupies an independent line. Table columns describe sequence and tissue related parameters, such as tissue fractions, TR, TE, FA, the measured signal, and tissue-type dependent relaxation constants. Default parameter values can be added using the `SetDefaultValues` methods. Absolute quantification^40–43^ is carried out via the `AbsQuant` method, using water as a reference and handling tissue heterogeneity and relaxation corrections.

### Auxiliary Classes and Visualization Tools

VDI offers several auxiliary classes, designed to facilitate loading, processing and display of data. Examples below will illustrate the use of these classes in real-life applications; here, we highlight the major ones.

`VDIIO` is used to read and write common data formats, including NifTI (.nii), DICOM, Bruker Paravision 5 and 360, and Siemens propriety (.rda) formats. It consists mostly of static methods – that is, methods which do not require a specific instantiation of a `VDIIO` object. Some of the most useful ones are `LoadNII` and `WriteNII` (reading and writing NifTI files into/from `VDIImage` objects), `LoadDICOMImageSeries` (reading a DICOM image series) and `LoadTwix` (reading Siemens raw data as `VDIImageND` objects).

`VDIPlot` contains multiple routines for efficiently partitioning tabular data and producing bar, line and correlation plots, such as `PlotTableBars`, Plot. This can be used to plot the results of absolute quantification. For example, `PlotTableBars(myTable, ‘conc’, {‘metabName’, ‘ROI’})` partitions the data in the MATLAB table `myTable` according to the different values in the variable columns `metabName` and `ROI` (region-of-interest), calculates the mean and standard-error-of-the-mean of the data stored in the variable `conc` within each such sub-partition, and produces a bar plot of these quantities.

The ability to interactively view, modify and process MRI and MRSI data is an essential component of scientific discovery. VDI includes a class, `VDIImageBrowser`, specifically designed for the visualization of data in `VDIImage`, `VDIShape` and `VDIImageND` objects. VDIImageBrowser allows for exploring n-dimensional data by reslicing it (using real-time nearest-neighbors interpolation) and overlaying different datasets, binary masks, and shapes in a common space.

## Methods

### Preprocessing and Fitting SemiLASER Data

Our first example script loads a collection of Siemens raw data files (.dat) containing SemiLASER data from the dorsal anterior cingulate of ten healthy volunteers (TR/TE=7000/42 ms), and performing coil combination, trimming of pre-echo points in the FID, registration of each transient via alignment and phasing, averaging, Fourier transformation and automated phasing. The script then creates a basis set by an idealized SemiLASER sequence with idealized hard pulses, and proceeds to fit it using the LCModel package. Default fitting parameters are used, except the `dkntmn` parameter which controls the baseline tortuosity (almost all LCModel control file parameter can be passed to the `FitLCModel` method). Finally, it carries out absolute quantification. Data were acquired on a 7T Siemens Terra MRI scanner, using the default single-channel transmit/24 channel receive Nova coil; the data were been previously published as part of a comparative study between edited and unedited quantification at ultrahigh fields^44^.

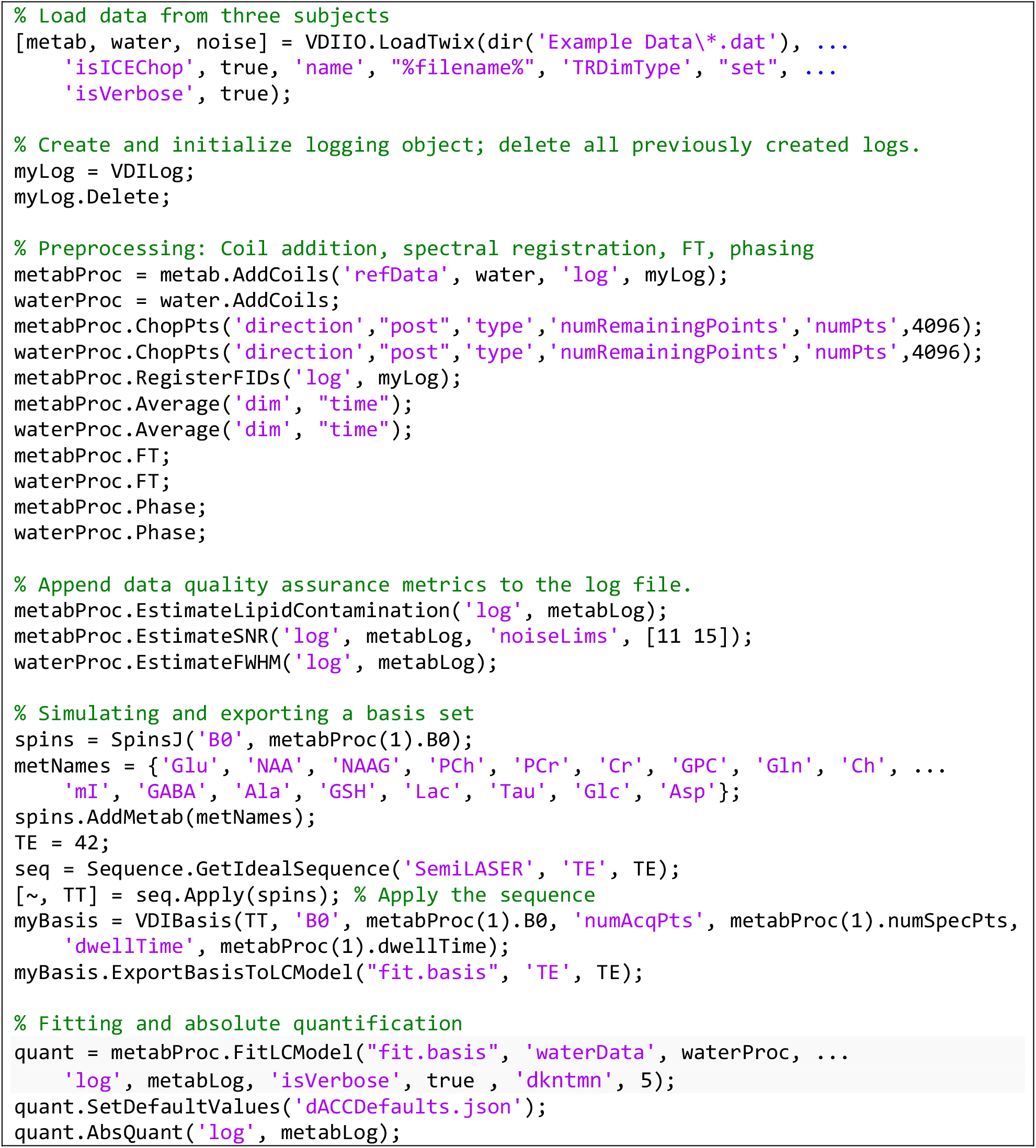

The static method LoadTwix in the `VDIIO` class is used to read all raw Siemens data (*.dat) in the Example Data directory. Data was acquired using a widely available SemiLASER implementation by the CMRR spectroscopy group (https://www.cmrr.umn.edu/spectro/); Setting the `isICEChop` option to true removes the pre-echo points acquired by the sequences (as per information stored in the ICE Siemens header), and the name variable tells the function how to populate the name property of each loaded dataset. Almost all methods accept an `isVerbose` parameter, providing real time processing feedback to the command line as the methods are executed. `LoadTwix` returns arrays of `VDIImageND` objects containing metabolite, reference water and noise data from each of the ten volunteers scanned: `metab(k)` is the `VDIImageND` object corresponding to the metabolite data from the k^th^ volunteer. The subsequent lines perform preprocessing of both the water and metabolite data. Nearly all methods accept a `VDILog` object via the log input parameter, and output some form of feedback to the HTML report. The `VDILog` object is initialized before preprocessing takes place, and deletes the previous report file.

Basis simulation follows using the `SpinsJ` and `Sequence` classes. A coupled spin structure is defined and populated with fifteen metabolites, starting out at thermal equilibrium. An idealized SemiLASER sequence using hard pulses is initialized, and applied to the spins. The resulting `TransitionTable` object, `TT`, is fed into a `VDIBasis` object with the appropriate spectral parameters, which is used to create a basis file in a format readable by LCModel.

Once a basis set is generated, fitting can take place using the publicly-available LCModel package via the `FitLCModel` method. This method only requires the name of an appropriate basis file; it obtains all required metadata from the basis file or from the `VDIImageND` object which calls it. It operates sequentially on all loaded single-voxel datasets in the 1×10 array `metabProc`. Additional LCModel parameters can be passed via additional input switches to FitLCModel; here, the baseline is constrained to remain linear by setting the LCModel `dkntmn` parameter to five. `FitLCModel` not only saves all processed control and output files generated by LCModel, but also reads them and returns a `VDIQuant` object, `quant`, which is subsequently used for absolute quantification^42,43,45–47^. `quant` is populated with metadata from the `VDIImageND` objects; however, a few parameters which are not stored in these objects – such as the region of interest, tissue fractions, transmit inhomogeneity and tissue relaxation values – default to values stored in the file `dACCDefaults.json`. JavaScript Object Notation (JSON) is an open standard file format, widely adopted by is which is a short text file containing default values for parameters not found elsewhere:

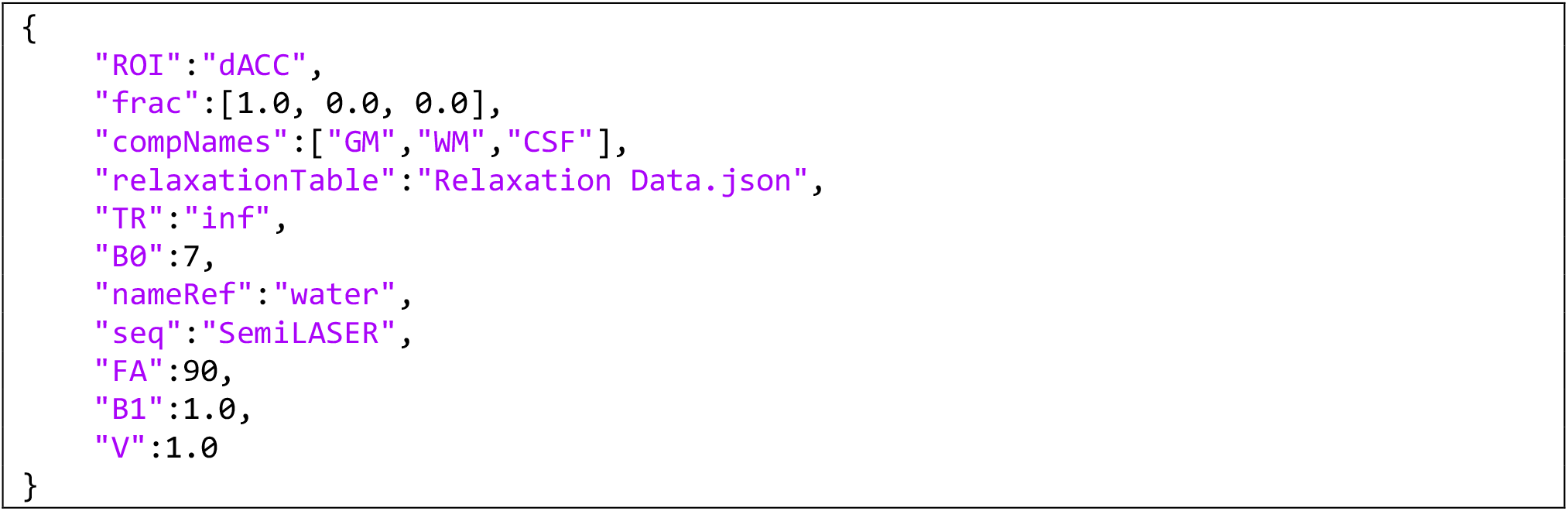

Most importantly, this file points towards another a second JSON file, `Relaxation Data.json`, containing relaxation data for metabolites and reference water from multiple ROIs, summarized from the literature. Because of the paucity of relaxometry studies at the dACC at 7T, the T_1_ and T_2_ values used herein originate from other regions with similar GM and WM composition.

### Global Metabolite Concentrations Using Linear Regression

Global linear regression extracts tissue-specific metabolite concentrations over the entire MRSI VOI. Such an approach is superior to ROI-based approaches for assessing diffuse disease activity^48^, because it circumvents two major problems with MRSI: The poor SNR in each voxel, and the partial volume due to a mixture of both GM, WM and CSF tissue in each voxel. Briefly, linear regression models the signal in each voxel as a linear combination arising from each of the voxels, assuming a global, single concentration for each tissue type (Fig. 2): 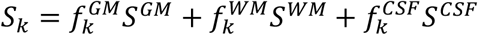, where k is the voxel index, and 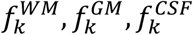 are the tissue fractions 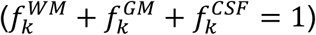. Since no detectable metabolite signals are observed in the CSF, *S*^*CSF*^ = 0. This approach is discussed in depth in the literature^21^, and has been used extensively to study healthy physiology^49–51^, diffuse axonal injury in mild traumatic brain injury^23,48,52–57^, and normal appearing white matter in multiple sclerosis^22,58^. Recently, linear regression was also adapted to sodium MRI^59^.

**Figure 2.**
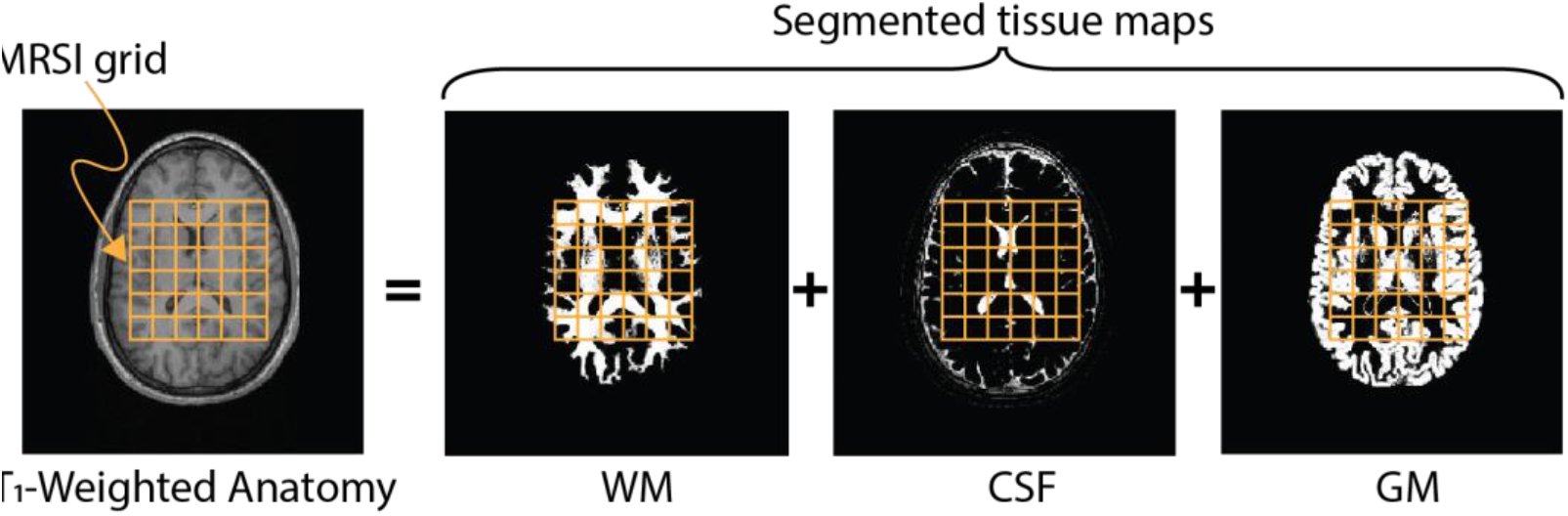
In linear tissue regression, the MRSI metabolite’s signal in each voxel is modeled as a linear combination of the contributions of each tissue type (GM, WM, CSF), proportional to the amount of each tissue in each voxel.

The application of linear regression to MRSI data becomes straightforward with VDI, which interfaces with SPM to generate tissue maps from high resolution T_1_-weighted anatomical images. The required VDI code is:

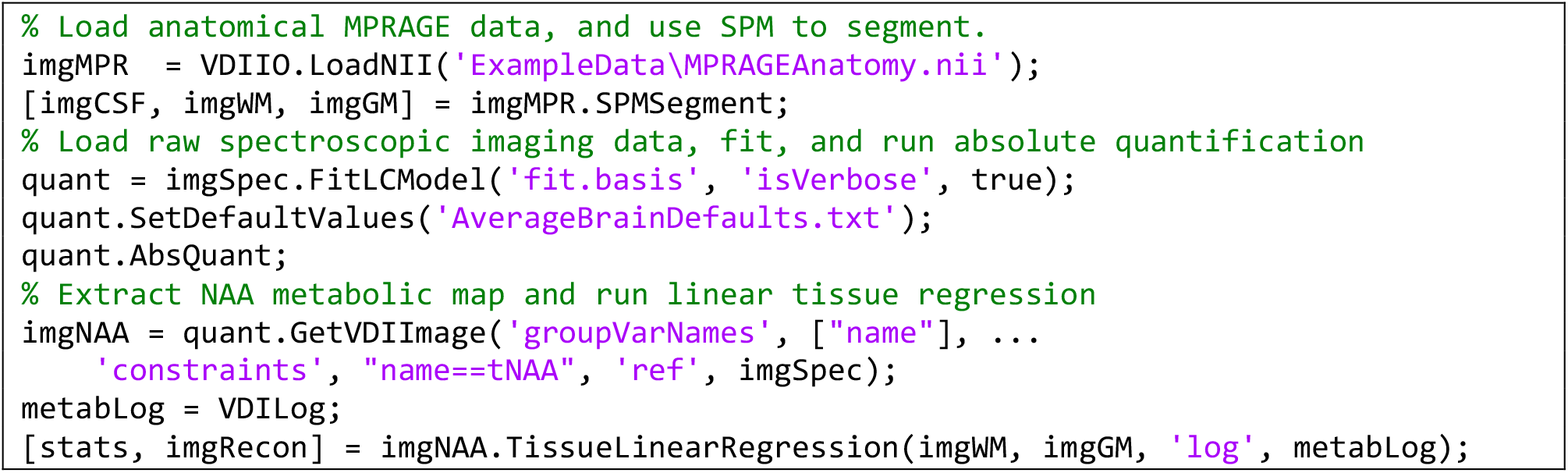

The first two lines load an anatomical T_1_-weighted MPRAGE image and uses SPM12 (added to the MATLAB path) to segment it into three `VDIImage` high resolution tissue maps. A previously loaded `VDIImageND` object, `imgSpec`, containing MRSI data from a 2D slice with a 32×32 in-plane matrix is fit on a voxel-by-voxel basis using LCModel; this step is time consuming, since each voxel takes about 10 seconds to fit, requiring a total processing time of approximately 32×32×10 seconds≈3-4 hours. As with the single-voxel example, it returns a `VDIQuant` object (quant), which contains fitted metabolite signals from each and every voxel. This undergoes fitting using an analogous pipeline to the single-voxel case. The `GetVDIImage` method of the `VDIQuant` class is then used to create `imgNAA`, a metabolic map of total NAA of class `VDIImage`, containing the concentration of NAA, and using the spatial orientation and positioning of the original `imgSpec`. Finally, linear tissue regression is run via the final line, and its results are both returned as variables and appended to an existing log file.

### Metabolite Concentrations from Atlas-Defined ROIs

When working with an anatomical binary mask (a `VDIImage` object) and an MRSI dataset (a `VDIImageND` object), VDI offers a convenient method called `CalcStats` to calculate metabolite averages, medians, and other statistics. Automation is possible when an anatomical high-resolution T_1_-weighted image is available. In such cases, the image can be registered to a common space like MNI, and the inverse transformation can be applied alongside a predefined atlas to generate anatomical masks in the subject’s original space. VDI simplifies the handling of these transformations through its `SPMGetAtlasStructure` method, which automates the relevant SPM routines. Atlases are comprised of two files: a NifTI image containing imaging data in a common space (e.g., MNI), and an XML file that encodes various atlas labels, such as those found in the Automated Anatomical Labeling (AAL) atlas^60^. To use these atlases with VDI, the corresponding files should be placed in the VDI atlases subdirectory. This enables VDI to not only detect their presence but also provide auto-completion for all available label names.

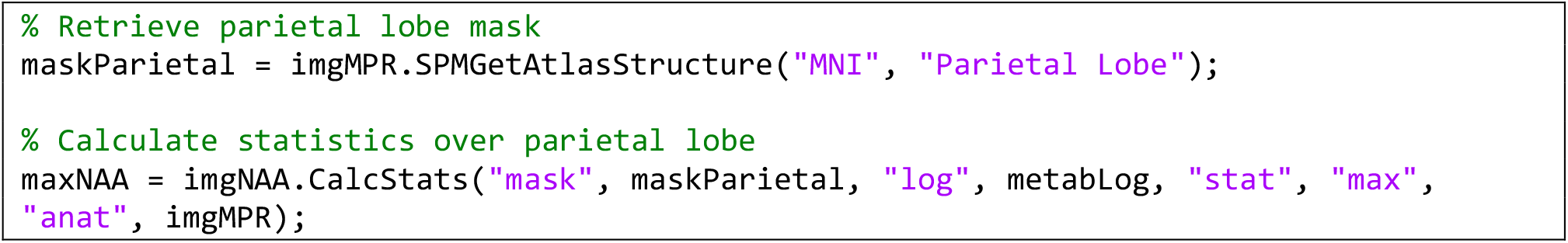

## Results

### Preprocessing and Fitting SemiLASER Data

Fig. 3 shows the output of several of the preprocessing steps, as well as fitting and quantification. All plots were taken directly from the generated log file. The spectral registration (Fig. 3A) plots the mean spectrum, taken over all ten volunteers (dark blue), and the spectra from each individual volunteer (light blue), before and after spectral registration. The relative stability of the magnet is visually reflected by the similarity between the spectra before and after registration. A more quantitative view can be had by looking at the zero order phases and spectral shifts of each FID, shown in the next two plots, for all subjects. As expected, greater shifts are required for later FIDs, as the magnet drifts slightly over time. Automatic phasing is shown in Fig. 3B for all subjects before and after phasing, as well as for a sample subject. A histogram showing the distribution of zero order phases is provided, showing the mean zero-order phase was 36°, with some inter-subject variability (standard deviation: 16°). The third line (Fig. 3C) shows quality assurance plots: lipid contamination, water FWHM (in ppm) and SNR taken over all volunteers, as well as an image showing the spectroscopic signal from the region designated as noise. This image provides visual reassurance that the region does indeed contain normally distributed noise; a small, linear bias can be seen, likely due to the slight imperfections in the phasing of the residual water baseline. Finally, the mean spectrum, its standard deviation, the mean fit and standard deviation, residual and baseline, over all volunteers, are shown in Fig. 3D, alongside a bar plot showing the quantified concentrations from the voxel.

**Figure 3.**
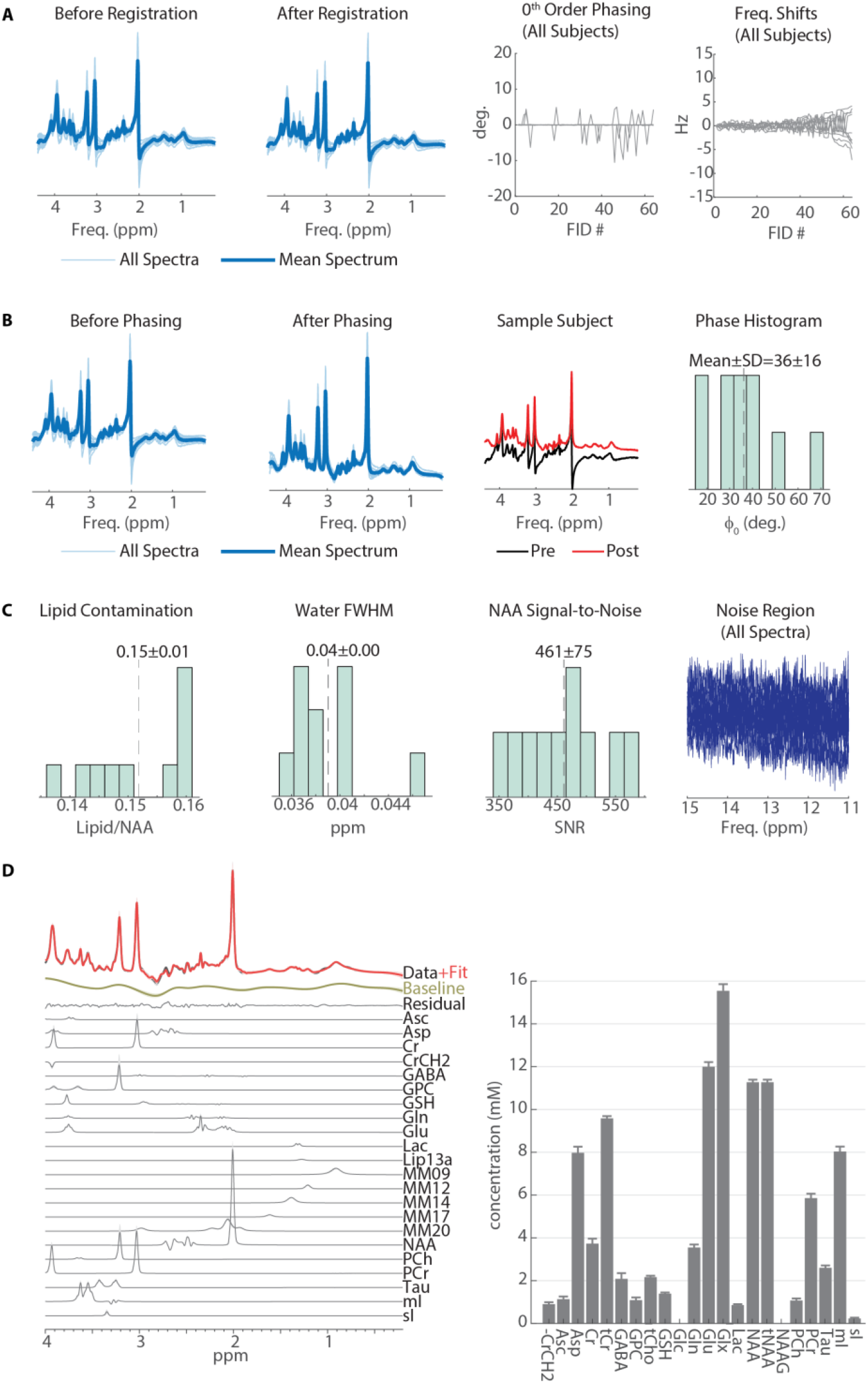
Spectral preprocessing and fitting of SemiLASER data acquired at 7T from 10 volunteers. All figures were taken with minor modifications from the logging files produced by the MATLAB scripts using VDI (via the VDILog library). (A) Spectral registration. (B) Zero order automated minimum entropy phasing. (C) Quality assurance metrics, including lipid contamination (relative to NAA), water full-width-at-half-maximum, NAA SNR, and a visualization of the region used to estimate noise. (D) Results of spectral fitting and absolute quantification.

### Tissue and Region Specific Metabolite Concentrations

Fig. 4 showcases the results of the linear regression for obtaining mean tissue-type concentrations. As with the single-voxel analysis, all plots and details shown are taken directly from the output log file. A total of 248 voxels out of the full 32×32=1024 in-plane 2D matrix were used; these were chosen as those voxels for which the combined WM and GM fractions exceeded 80%. The mean tissue concentrations were 11.50 mM (95% confidence interval: [11.08, 11.93] mM) for GM, and 10.54 mM (95% confidence interval: [10.01, 11.06] mM) for WM, with R^2^=0.156 (Fig. 4A). The variance inflation factor of GM and WM fractions vectors was 0.35, indicating the two regressors do not exhibit substantial co-linearity. Histograms of GM, WM fractions for the 248 voxels are shown in Fig. 4B, and the signal vs. the WM fraction is plotted in Fig. 4C, indicating voxel tNAA concentrations decrease with increasing WM content, as expected from the literature^21^. Fig. 4D shows the residuals obtained by subtracting the fitted linear model values from the quantified concentrations in each voxel. Figs. 4E-G show the map of tNAA, the spatial mask of the voxels used in the regression analysis, and the linear reconstructed tNAA map. Finally, Fig. 4H shows the map of residuals, the values of which are shown in the histogram in Fig. 4D.

**Figure 4.**
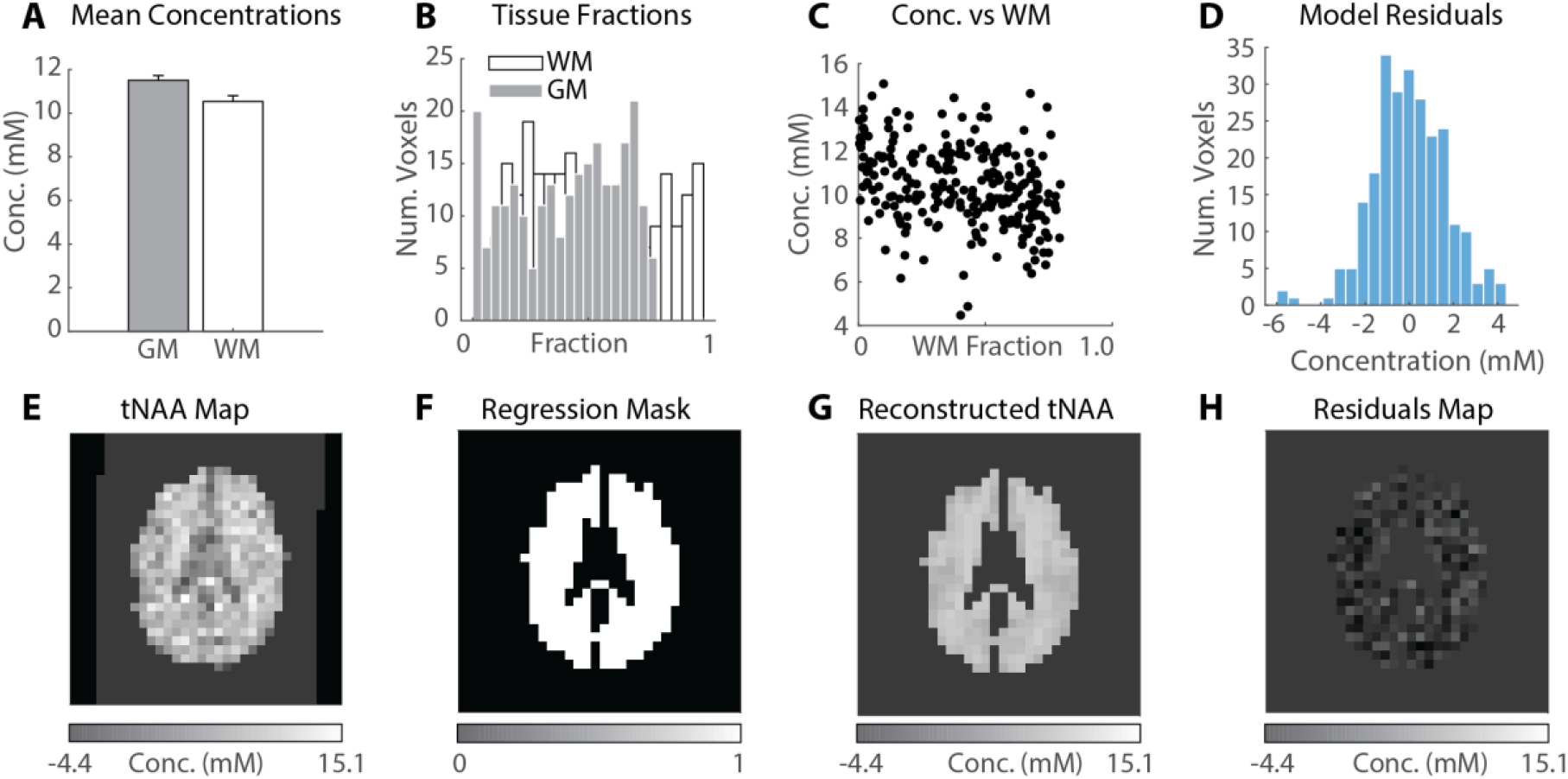
Linear tissue regression. (A) Mean±standard error of the global NAA concentration in each tissue type. (B) Histogram sof gray and white matter fractions in the spectroscopic voxels used in the regression. (C) NAA concentrations as a function of white matter fractions (each point is a separate spectroscopic voxel). (D) Histogram of fitting residuals, i.e. the difference between data and fitted values in each voxel. Note the distribution is normal, as should be. (E) Metabolic map of NAA (stored in the `imgNAA` variable). (F) Binary mask over which regression is carried out, defined by the brain segmentation and the requirement WM+GM≥0.8. (G) Reconstructed NAA map obtained from linear regression. (H) Spatial map of residuals (same numbers used in (D)). All metabolic maps employ the same intensity scale for consistency, but only the residuals exhibit negative values.

Fig. 5 shows the parietal lobe that was automatically generated from the MNI atlas overlaid on top of both the metabolic tNAA map and the reference MPRAGE image, as well as the histogram of tNAA concentrations from the mask voxels. A total of 47 voxels fall within the mask, with a mean±standard deviation concentration of 8.83±2.02 mM and a range of [2.31, 10.95] mM.

**Figure 5.**
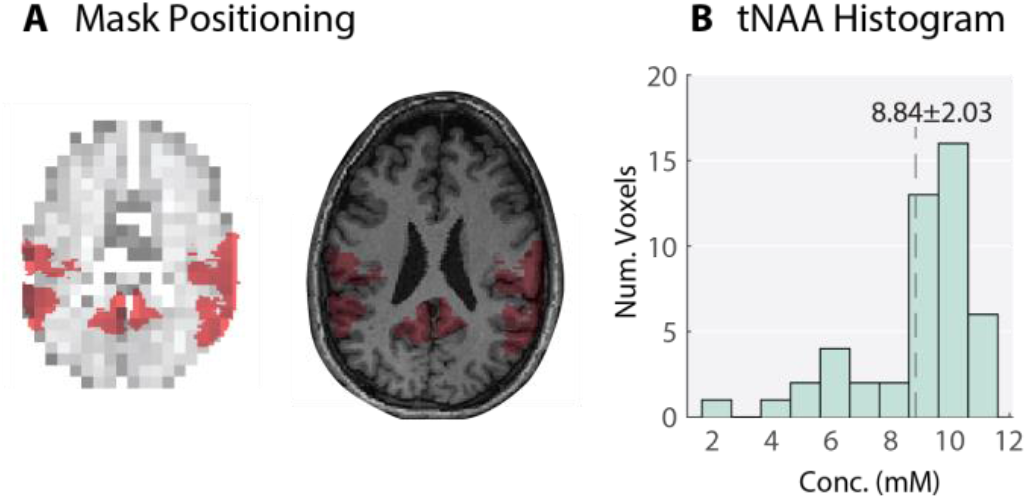
ROI tNAA concentrations from a predefined mask (parietal lobe). (A) Mask localization on both the metabolic map and anatomical MPRAGE image. (B) Histogram of tNAA concentrations from the mask.

## Discussion

The increasing complexity and size of neuroimaging datasets requires tools that are not merely efficient and accurate, but also easy to use, self-documenting and equipped with the ability to alert the user about potential pitfalls. For example, the recent focus on harmonization techniques of large-scale multisite MRS data is one such indication of this trend^15,61,62^. VDI was constructed to address such a need; It focuses on providing its users with a modern, extensively-documented object-oriented framework that promotes writing self-documenting code and minimizes tedious tasks. Its methods were written to promote batch processing of data, as shown in the first example where several single-voxel datasets were processed simultaneously.

A second major focus of VDI is on its automated reporting capabilities. Succinct code often runs the risk of hiding assumptions, and complex processing pipelines could fail in surprising ways while deceptively outputting seemingly-meaningful numbers. Both challenges can be overcome by detailed reporting capabilities. VDI implements this philosophy by making it possible to generate HTML log files. These provide detailed reports of the parameters used, even those not directly input into each method, as well as visual feedback, quickly indicating whether a particular processing step has failed. All figures generated are saved by default both as bitmaps and vector graphics, making them eminently usable in scientific publications, as has been done herein (Figs. 3, 4). In addition, each processing step attempts to automatically identify failures and outliers and alert the users to their presence at the appropriate place in the report. For example, when phasing multiple datasets simultaneously, individual datasets which significantly differ in their zero order phase from the group median by more than three standard deviations are listed in red in the report.

Unlike many other MRS packages, VDI does not offer a graphical user interface, except when visualizing raw MRS data. While graphical interfaces appeal to newcomers, they are prone to manual user errors which are difficult to identify and correct. The underlying philosophy of VDI instead focuses on writing reproducible, self-documenting scripts, and instead relegates visual feedback to the reporting stage, after all processing has ended.

MATLAB remains a widely used, popular platform for scientific computing and data analysis, available in many major universities and academic institutions. Although MATLAB itself is a commercial product, VDI does not require any additional toolboxes to operate, and is free and open source, made available through a modern version control framework (GitHub). It also makes use of standardized file formats for storing and using metadata, such as JSON and XML formats for relaxation data and atlas label definitions, in an attempt to increase standardization among the MRS community. Such efforts are becoming increasingly common, as evident by a range of consensus papers on its methodology, analysis and reporting standards^42,63–68^.

## Data Availability

The VDI software and all example data discussed herein are available from the software website directly, at www.vdisoftware.net

## Software Availability

VDI can be downloaded from the corresponding Author’s website: https://www.vdisoftware.net

## Notes

**Sponsors, Grants & Funding:** Assaf Tal acknowledges the support of the Monroy-Marks Career Development Fund and the historic generosity of the Harold Perlman Family. Assaf Tal acknowledges the support of the Israel Science Foundation (Personal Grant 416/20). Yiling Liu and Zhiyong Zhang acknowledge the support of the National Natural Science Foundation of China (No. 62001290)

### Competing Interest Statement

The authors have declared no competing interest.

### Funding Statement

the Israel Science Foundation (Personal Grant 416/20); the National Natural Science Foundation of China (Grant No. 62001290).

### Author Declarations

All data used in the current publication were previously published (T. Finkelman, E. Furman-Haran, R. Paz and A. Tal, NeuroImage 247(15):118810 (2022)), made available publicly by request from Authors, and approved by an appropriate IRB board.

